# Investigating of the causes of seizures in children admitted to Kerman hospitals in 2017

**DOI:** 10.1101/2023.06.28.23292005

**Authors:** Hossein Ghaedamini, Habibe Nejad Biglari, Zahra Saghafi, Ali Hosseininasab, Ali Amirbeigi, Farzad Ahmadabadi, Amirmahdi Ghaedamini

**Affiliations:** Department of Surgery diseases, School of Medicine, Golestan Hospital, Jundishapur University of Medical Sciences, Ahvaz, Iran; Kerman University of Medical Sciences, Kerman, Iran; Getriatric Care Research Center, Rafsanjan University of Medical Sciences, Rafsanjan, Iran; Peadiartic Department, Nursing and Midwifery School, Rafsanjan University Of Medical Sciences, Rafsanjan, Iran; Ardabil University of Medical Sciences, Ardabil, Iran; Research Committee of Medical school, Tehran University of Medical Sciences, Tehran, Iran

**Keywords:** Seizure, Children, Etiology

## Abstract

**Introduction:** Seizure in children with the prevalence of 4 to 6 cases in 1000 is the most common neurological disorder in pediatrics. This study aimed to determine the causes of seizures in children admitted to Kerman hospitals in 2017

**Materials and methods:** In this retrospective study, 250 hospitalized children suffering from seizure were selected through census method. Data collection tool was a checklist. The obtained data was analyzed employing SPSS_24_ software, using chi-square, and correlation coefficient statistical tests at the significant level P < 0.05.

**Results:** Out of 250 children studied, 55.6%were male and 44.4%were female. The mean age of the children was 10.54 ± 3.7 years. 80.4%were born with vaginal delivery and 19.6%were born with cesarean section. 44%of the patients had febrile seizure, 23.2%epilepsy, 12.4%infection, 10%gastroenteritis, 5.2%static encephalopathy, 2.8%meningitis, 1.6%hypocalcaemia, 0.4 %metabolic disorders and 0.4 %tumor. That underlying factors of febrile seizure were upper respiratory infection (43.6%), gastroenteritis (15.7%), shigellosis (10.5%), urinary infection (9.9%), pneumonia (8.1%), unidentified fever (7.8%), otitis (3%) and dental abscess (1.4%) respectively.

**Conclusion:** According to this study, the most common cause of the seizure is a febrile seizure, which confirms the results of other studies. Common seizure require diagnosis, planning, and special treatments that can be obtained by taking a complete history, accurate examination, and effective Para clinical evaluations.

## Introduction

Seizures a transient occurrence of signs and/or symptoms due to abnormal excessive or synchronous neuronal activity in the brain (1). They happen because of sudden, abnormal electrical activity in the brain (2,3). Five million people in the world have seizures (three million of them are children) (4). The results showed that seizures in children have the prevalence of 4 to 6 cases in 1000 people and it is the most common neurological disorder in pediatrics. In the last five years, about 880,000 children and adolescents in the United States need medical attention for evaluation of a newly occurring seizure disorder (5). The prevalence of seizures in Iran was estimated to be around 5%(6). There is a large difference in the incidence rate of Febrile Seizures (FS) around the world. Between five percent of all children less than five years old in Europe and the United States experience at basically the least one seizure associated with a febrile illness (7).

The study of Delpisheh and et al conducted in Iran (4599 people) showed that the prevalence of FS according to the age of children under 2 years and 2 to 6 years were 55.8%and 44.1%, respectively in Iran (8). Seizures can caused by chemicals in the blood, infections like meningitis or encephalitis (10, 11). A common cause of seizure is fevers (12). Other causes include head trauma, stroke or lack of oxygen to the brain, genetic defects or brain tumors, hypoglycemia and deficiency of vitamin B6 (13-16)

With the epidemiological studies in the field of children’s seizure disorder, the incidence, prevalence, genetic background, age, and therapeutic approaches can be evaluated (17).

In a 14-year follow up (in the felid of neurological complications of fibrillary seizure), the results showed that 17%of children had the neurodegenerative developmental delay, 10%had neurological anomalies, and 4%had neurological defects (18).

The study of Abbaskhanian and et al. Showed that prevalence of neonatal seizure was 12.4%in Sari city (north of Iran) (19). The results of Sabzehei and et al showed that the prevelance of seizures was 9.1 %(20).

On the other hand, the diagnosis of seizures in children is one of the major problems for pediatric residents and general predictions (20). It takes a lot of time and energy (19).

Due to high prevalence of seizures, its psychosocial and cognitive importance, and determining the underlying factors of the disorder, in addition no study evaluated the frequency of seizures in the southeastern region of Iran (New Aspect) and other existing studies are old, so the present study was conducted to determine the causes of seizures in children admitted to Kerman hospitals in 2017

## Materials and methods

This retrospective cross-sectional study was conducted on all children aged 1 month to 16 years with the initial diagnosis of Seizure that admitted to Kerman hospitals (included Afzalipour, Sahid Bahonar, and Shafa) from January to September 2017.

Kerman is located in the south-central part of Iran. On December 31, 2016, the population in ages from 1 month up to 16 years was 135,688 people (21). There were 65,147 girls and 70,541 boys.

This research was performed using the hospital data registration medical records in order to select all children with the diagnosis ‘Seizure’ recognized between January to September 2017 (22).

The Statistical Population was all children aged 1 month to 16 years with the initial diagnosis of Seizure that admitted to Kerman hospitals (included Afzalipour, Sahid Bahonar, and Shafa) from January to September 2017.

Sampling was done using the census method (Sample Size= 250 Participants) and the sample size was all children aged 1 month to 16 years due to abnormal movements or abnormal behavior admitted to Kerman Hospitals with a diagnosis of Seizure.

Data collection was done through a checklist (data collection form) which included demographic data, family history, age at seizure onset, type of seizure, perinatal data, mental and motor development, seizure types, other diseases accompanied with seizures as a symptom, treatment.

### Definition

Seizure defined as is a period of symptoms due to abnormally excessive or synchronous neuronal activity in the brain. (23-25).

## Diagnosis and classification

Diagnosis of seizure and classification of etiology of seizure in children should be based on history, physical examination, and clinical judgment of one pediatric neurologist based on ILAE (International League against Epilepsy) criteria (26).

## Statistical methods and statistical tests

We used mean and standard deviation and relative and absolute frequency to analyze quantitative and qualitative variables, respectively. Moreover, to investigate the association between demographic and epidemiological factors of seizure types, χ ² and Fisher’s Exact Test was performed. Then, the obtained data was analyzed employing Statistical package for particularly social science (SPSS) version 24 (IBM Inc., Chicago, IL, USA) and correlation coefficient statistical tests at the significant level P < 0.05.

## Ethical considerations

The research protocol was approved in the Medical Ethics Committee of Kerman University of Medical Sciences (Ethical Code: IR.KMU.AH.REC.1395.659).

## Results

55.6%of participants (total participants= 250) were male and 44.4%were female. 80.4%born with Normal Vaginal Delivery (NVD) and 19.6%were born with Cesarean Section (C/S). The mean ± SD of the duration of admission was 8.73 ± 1.2 days, the mean ± SD age of the children was 10.54 ± 3.7 years, and the mean ± SD birth weight was 2675.48gr ± 453.64, 40.4%of children had a seizure for the first time. The frequency of seizure repetition in 35.1%of the children was 4 times a day and 38.5%of them had a positive familial history. Table 1 shows seizures causes. The most common cause was a febrile seizure (45.5%).

**Table 1.** The seizure’s causes among admitted Children in Kerman hospitals.

| Etiology | Seizure |  |
| --- | --- | --- |
|  | Frequency | Percentage |
| Febrile Seizure | 110 | 44 |
| Epilepsy | 58 | 23.2 |
| Infection | 31 | 12.4 |
| Gastroenteritis | 25 | 10 |
| Static Encephalopathy | 13 | 5.2 |
| Meningitis | 7 | 2.8 |
| Hypocalcaemia | 4 | 1.6 |
| Metabolic Disorders | 1 | 0.4 |
| Tumor | 1 | 0.4 |
| Total | 250 | 100 |

There was not any significant association between gender (P=0.35), type of delivery (P=0.83), birth weight (P=0.56) with causes of seizures but there was a significant association between age at the onset of seizures with it (P<0.01).

The result showed that underlying factors of febrile seizure were upper respiratory infection (43.6%), gastroenteritis (15.7%), shigellosis (10.5%), urinary infection (9.9%), pneumonia (8.1%), unidentified fever (7.8%), otitis (3%) and dental abscess (1.4%) respectively (Table 2).

**Table 2.** The frequency of febrile seizure’s etiology among admitted Children in hospitals of Kerman:

| Name of disease | Frequency | Percentage |
| --- | --- | --- |
| Upper Respiratory Infection | 46 | 41.8 |
| Gastroenteritis | 19 | 17.2 |
| Shigellosis | 14 | 12.8 |
| Urinary Infection | 10 | 9 |
| Pneumonia | 9 | 8.2 |
| Unidentified Fever | 7 | 6.4 |
| Otitis | 4 | 3.7 |
| Dental Abscess | 1 | 0.9 |
| Total | 110 | 100 |

Furthermore, the most common types of seizures were focal with Secondary generalization (25.6%), focal (16.4%), myoclonic (12%), and atonic (11.6%) respectively (Table 3).

**Table 3.**
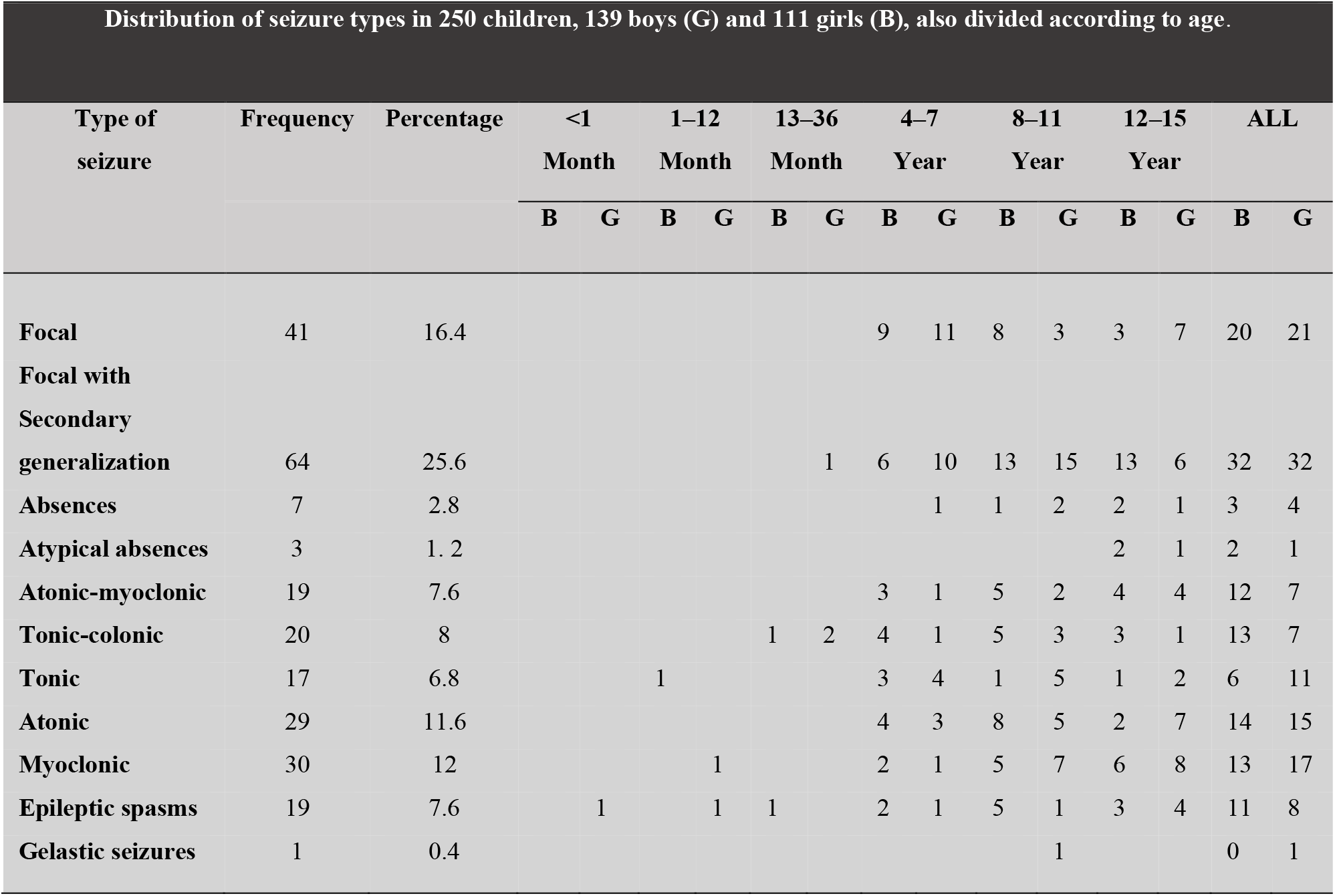
Distribution of seizure types in 250 children, 139 boys (G) and 111 girls (B), also divided according to age.

## Discussion

Considering the cultural and socioeconomic differences, food habits, and regional diseases, this study was focused on the causes of children’s seizures in patients admitted to hospitals of Kerman in order to find out its causes and resolve them and finally prevent the occurrence of seizure attacks.

This was the first study in the southeastern region of the country to investigate the causes of seizures in children, which is important.

In our study, most of the children’s seizures happened in the boys, which is similar to previous studies (27-25). The cause of it is controversial and needs complementary investigations.

On the other hand, it is probable that more attention is paid to boys in developing countries lead to the number of identified boys be higher than girls (30, 31).

The prevalence of FS in this study is relatively high compared with other studies, which is due to the fact that patients are referred from nearby provinces and other cities to Kerman hospitals (32).

The common causes of seizures were FS, epilepsy, infection, gastroenteritis, static encephalopathy, meningitis, hypocalcemia, metabolic disorders, and tumor which is similar to many studies (30, 33).

The most common cause of seizure was FS (44%) that is similar to many studies (27-33). Proper warnings and suitable training should be given to parents who have a child with a history of FS (34). Therefore, rectal diazepam should be given for prophylaxis in these children when fever is higher than 38 degrees of centigrade (30).

The second cause of seizure was epilepsy (23.2 %) which is similar to many studies (30, 33). Epilepsy is a common disorder in developed countries (about two to four percent) which is higher in developing countries (35).

The third cause of seizure was Infection (12.4 %) that is similar to many studies (30, 33). Infections are a major cause of seizures in developing countries like Iran (36). It can cause both acute symptomatic seizures and epilepsy (37). The etiologies of Infectious include tuberculosis, HIV, cerebral malaria, neurocysticercosis, subacute sclerosing panencephalitis, cerebral toxoplasmosis that sometimes have a structural correlate (35).

The next cause of the seizure was gastroenteritis (17.2 %). Seizures due to gastroenteritis are seen in infants and children and known among neurologists and pediatricians which is similar to the results of this study (38).

On the other hand, shigellosis in children of developing countries is associated with high fever and seizures (39). In the study of Ashkenazi and colleagues, 153 children had shigellosis and seizures. 20%of them had a positive family history of seizure, indicating an important role of genetics in seizure due to gastroenteritis (40).

Despite other studies, the high frequency of seizures due to static encephalopathy (5.2 %) in our research can be attributed to poisoning, drug abuse, and mitochondrial disorders. The results showed that Meningitis (2.8 %) known as a cause of the seizure that is similar to other studies (41-43).

Therefore, if a child less than two years old had a seizure, meningitis could be one of the diagnoses, and more testing and examination should be done for r/o it (44). The results showed that the frequency of seizures due to hypocalcemia is 1.6 %and is related to the onset age of rickets symptoms in children (45).

Seizures due to metabolic disorders (0.4 %in our study) could be a presenting feature of late-onset metachromatic leukodystrophy caused due to lack of saposin B and arylsulfatase A enzyme deficiency (46).

The results showed that seizure can be one of the important symptoms of brain tumors that is similar to the results of other studies (47-49).

In this study, 94 children had previous history of seizure and 156 children (62.4%) was new cases, that indicating the important role of genetic, that is similar to textbooks of neurology (48). totally, 19%of children had a family history of febrile seizures that 35 participants were in the group with the FS diagnosis that is also similar to textbooks. Genetics not only affects the incidence of FS; It also affects the recurrence and type of it, as the probability of recurrence FS and complex seizures is higher in children with a positive family history (49).

From 250 children, 64 patients were treated with anticonvulsant medications that 19 people discontinued their medication during the last days and week’s which play an important role in the occurrence of recurrent seizures (50, 51).

Totally, the causes of children’s seizures in the Merritt’s textbook (52) include idiopathic epilepsy (3.5%), degenerative diseases (5.2%), infections (5.65%), malignancies (4.1%), trauma (5%), maternal diseases (8%) and vascular diseases (10.9%), which is different from the results of our study. The variation in results is probably explained by the location of the studies. Because this research conducted in a deprived province, and people of its community should be followed up until complete treatment. Also in these areas, people visit a doctor when their children get seizures, and they continue the treatment just for a short time. Some people do not pay enough attention to the seizures risk factors and medical advice and still insist on their own habits and traditions. Sometimes the high cost of treatment prevents patients from complementary follow-ups.

In this study, underlying factors of FS were upper respiratory infection (43.6%), gastroenteritis (15.7%), shigellosis (10.5%), urinary infection (9.9%), pneumonia (8.1%), unidentified fever (7.8%), otitis (3%) and dental abscess (1.4%). Some studies showed that the risk of seizure in children less than 3 years who have a viral upper respiratory infection with positive family history is higher than others (51-54).

This study had limitations on sample size and time interval and we could not investigate the association of some variables like maturity (term or preterm) and history of admission with causes of seizure. We suggest that future multicenter studies be conducted in larger sample sizes and extended time intervals.

## Conclusion

Altogether the results showed that the most common cause of children’s seizures was FS, Epilepsy, and Infection. So appropriate policies should be made to reduce the rate of them. Due to the economic costs and complications of seizure in our society regular monitoring and follow-ups of seizure can reduce the mortality and morbidity of it.

Conducting similar studies in other regions of Iran is necessary and help to detect other causes of seizure and achieve more comprehensive and complete results. It is suggested that other studies investigate the association between other causes of the seizure (did not mention in our study) in children and variables such as age, gender, and season.

## Data Availability

All data produced in the present work are contained in the manuscript

## Acknowledgments

The authors would like to thank the personnel of Afzalipour, Sahid Bahonar and Shafa hospitals of Kerman for their cooperation in performing the present study.

## References

1. Ekanem EE, Fajola AO, Usman R, Ogbimi RN, Ikeagwu GO, Anidima TE, Etieh MN, Umejiego CN. Management of epilepsies at the community cottage hospital level in a developing environment. Nigerian Medical Journal: Journal of the Nigeria Medical Association. 2019 Jul;60(4):186.

2. Bryndziar T, Sedova P, Kramer NM, Mandrekar J, Mikulik R, Brown Jr RD, Klaas JP. Seizures following ischemic stroke: frequency of occurrence and impact on outcome in a long-term population-based study. Journal of Stroke and Cerebrovascular Diseases. 2016 Jan 1;25(1):150–6.

3. Buelow JM, Shafer P, Shinnar R, Austin J, Dewar S, Long L, O’Hara K, Santilli N. Perspectives on seizure clusters: gaps in lexicon, awareness, and treatment. Epilepsy & Behavior. 2016 Apr 1;57:16–22.

4. Conejo Moreno D, Rodriguez Fernandez C, Ruiz Ayucar de la Vega I, Ortiz Madinaveitia S, Hedrera Fernandez A, Maldonado Ruiz E, et al. [Para-infectious seizures: A retrospective multicentre study]. Anales de pediatria (Barcelona, Spain : 2003). 2016;85(6):300–4.

5. Englot DJ, Magill ST, Han SJ, Chang EF, Berger MS, McDermott MW. Seizures in supratentorial meningioma: a systematic review and meta-analysis. Journal of neurosurgery. 2016 Jun 1;124(6):1552–61.

6. Sayehmiri K, Tavan H, Sayehmiri F, Mohammadi I V Carson K. Prevalence of epilepsy in iran: a meta-analysis and systematic review. Iran J Child Neurol. 2014 Fall;8(4):9–17. PMID: 25657765; PMCID: PMC4307363.

7. Gavvala JR, Schuele SU. New-onset seizure in adults and adolescents: a review. Jama. 2016 Dec 27;316(24):2657–68.

8. Gersner R, Dhamne SC, Zangen A, Pascual-Leone A, Rotenberg A. Bursts of high-frequency repetitive transcranial magnetic stimulation (rTMS), together with lorazepam, suppress seizures in a rat kainate status epilepticus model. Epilepsy & Behavior. 2016 Sep 1;62:136–9.

9. Habibi M, Hart F, Bainbridge J. The impact of psychoactive drugs on seizures and antiepileptic drugs. Current neurology and neuroscience reports. 2016 Aug;16(8):1–0.

10. Packer RM, Volk HA. Epilepsy beyond seizures: a review of the impact of epilepsy and its comorbidities on health‐related quality of life in dogs. Veterinary Record. 2015 Sep;177(12):306–15.

11. Jiao J, Harreby KR, Sevcencu C, Jensen W. Optimal Vagus Nerve Stimulation Frequency for Suppression of Spike-and-Wave Seizures in Rats. Artificial organs. 2016;40(6):E120–7.

12. Lagunju IA, Oyinlade AO, Babatunde OD. Seizure-related injuries in children and adolescents with epilepsy. Epilepsy & behavior : E&B. 2016;54:131–4.

13. Lopim GM, Vannucci Campos D, Gomes da Silva S, de Almeida AA, Lent R, Cavalheiro EA, et al. Relationship between seizure frequency and number of neuronal and non-neuronal cells in the hippocampus throughout the life of rats with epilepsy. Brain research. 2016;1634:179–86.

14. Nicastro N, Assal F, Seeck M. From here to epilepsy: the risk of seizure in patients with Alzheimer’s disease. Epileptic disorders : international epilepsy journal with videotape. 2016;18(1):1–12.

15. Nicholas R, Magliozzi R, Campbell G, Mahad D, Reynolds R. Temporal lobe cortical pathology and inhibitory GABA interneuron cell loss are associated with seizures in multiple sclerosis. Multiple sclerosis (Houndmills, Basingstoke, England). 2016;22(1):25–35.

16. Singh A, Trevick S. The Epidemiology of Global Epilepsy. Neurologic clinics. 2016;34(4):837–47.

17. Sun Y, Zhang G, Zhang X, Yan X, Li L, Xu C, et al. Time-frequency analysis of intracranial EEG in patients with myoclonic seizures. Brain research. 2016;1652:119–26.

18. Szita B, Hidasi Z. [Psychogenic nonepileptic seizures: overview and implications for practice]. Orvosi hetilap. 2016;157(20):767–75.

19. Takeshita E. [Dystrophinopathy and Seizure]. Brain and nerve = Shinkei kenkyu no shinpo. 2016;68(2):128–36.

20. Us Centers For Disease C, Prevention Epilepsy P. About one-half of adults with active epilepsy and seizures have annual family incomes under $25,000: The 2010 and 2013 US National Health Interview Surveys. Epilepsy & behavior : E&B. 2016;58:33–4.

21. Fouladseresht H, Safiri S, Moqaddasi M, Razeghi MS, Bazargan N. Prevalence of food and airborne allergens in allergic patients in Kerman. Journal of Kermanshah University of Medical Sciences (J Kermanshah Univ Med Sci). 2014;18(4):234–41.

22. Mohamed C, Kissani N. Early seizures in acute stroke. The Pan African medical journal. 2015;20:136.

23. Zanzmera P, Menon RN, Karkare K, Soni H, Jagtap S, Radhakrishnan A. Epilepsy with myoclonic absences: Electroclinical characteristics in a distinctive pediatric epilepsy phenotype. Epilepsy & behavior : E&B. 2016;64(Pt A):242–7.

24. Kim HK, Chin BS. Clinical features of seizures in patients with human immunodeficiency virus infection. 2015;30(6):694–9.

25. Likhachev SA, Astapenko AV, Osos EL, Zmachynskaya OL, Gvishch TG. [Ecstatic seizures]. Zhurnal nevrologii i psikhiatrii imeni SS Korsakova. 2015;115(5):100–2.

26. Scheffer IE, French J, Hirsch E, Jain S, Mathern GW, Moshé SL, et al. Classification of the epilepsies: New concepts for discussion and debate—Special report of the ILAE Classification Task Force of the Commission for Classification and Terminology. Epilepsia Open. 2016;1(1-2):37–44.

27. Sadeghian A, Damghanian M, Shariati M. Neonatal seizures in a rural Iranian district hospital: etiologies, incidence and predicting factors. Acta Med Iran 2012; 50(11): 760–4. 12.

28. Mwaniki M, Mathenge A, Gwer S, Mturi N, Bauni E, Newton CR, et al. Neonatal seizures in a rural Kenyan District Hospital: aetiology, incidence and outcome of hospitalization. BMC Med 2010; 8: 16.

29. Kohelet D, Shochat R, Lusky A, Reichman B. Risk factors for neonatal seizures in very low birthweight infants: population-based survey. J Child Neurol 2004; 19(2): 123–8.

30. Dehdashtian M, Momen AA, Ziae T, Moradkhani SH. Evaluation of seizure etiology in convulsive neonates admitted to Imam Khomeini and Abozar hospitals of Ahvaz 2004-2007. Jundishapur Sci MedJ 2009; 8(2): 163–8.

31. Bhalla D, Lotfalinezhad E. A short perspective on the risk profile of epilepsy in Iran. Mathews Journal of Neurology. 2016;1(1):003.

32. Reinholdson J, Olsson I, Edelvik A, Hallbook T, Lundgren J, Rydenhag B, et al. Long-term follow-up after epilepsy surgery in infancy and early childhood--a prospective population based observational study. Seizure. 2015;30:83–9.

33. Riegle MA, Masicampo ML, Shan HQ, Xu V, Godwin DW. Ethosuximide Reduces Mortality and Seizure Severity in Response to Pentylenetetrazole Treatment During Ethanol Withdrawal. Alcohol and alcoholism (Oxford, Oxfordshire). 2015;50(5):501–8.

34. Staack AM, Steinhoff BJ. [Differential Diagnosis of Epileptic and Psychogenic Non-Epileptic Seizures and Treatment Consequences]. Fortschritte der Neurologie-Psychiatrie. 2015;83(12):702–35.

35. Herzog AG, Mandle HB, Cahill KE, Fowler KM, Hauser WA. Differential impact of contraceptive methods on seizures varies by antiepileptic drug category: Findings of the Epilepsy Birth Control Registry. Epilepsy & behavior : E&B. 2016;60:112–7.

36. Verdinelli C, Olsson I, Edelvik A, Hallbook T, Rydenhag B, Malmgren K. A longterm patient perspective after hemispherotomy--a population based study. Seizure. 2015;30:76–82.

37. Williams AM, Park SH. Seizure associated with clozapine: incidence, etiology, and management. CNS drugs. 2015;29(2):101–11.

38. Vafaei I, Rezazadehsaatlou M, Abdinia B, Khaneshi M, Hasanpour R, Panje F, et al. Study of the Determinant Factors in Seizure Following Gastroenteritis in Children Admitted in Tabriz Children’s Hospital during 2001 to 2016. International Journal of Pediatrics. 2017:6439–46.

39. Raju M, Kumar MP. Study of Association between Iron Deficiency Anaemia and Febrile Seizures. J Evol Med Dent Sci. 2015;4(39):6818–23.

40. Afroze F, Ahmed T, Sarmin M, SMSB Shahid A, Shahunja KM, Shahrin L, Chisti MJ. Risk factors and outcome of Shigella encephalopathy in Bangladeshi children. PLoS neglected tropical diseases. 2017 Apr 28;11(4):e0005561.

41. Aghaei-Lasboo A, Fisher RS. Methods for Measuring Seizure Frequency and Severity. Neurologic clinics. 2016;34(2):383–94.

42. Al Kasab S, Dawson RA, Jaramillo JL, Halford JJ. Correlation of seizure frequency and medication down-titration rate during video-EEG monitoring. Epilepsy & Behavior. 2016 Nov 1;64:51–6.

43. Amorim BO, Hamani C, Ferreira E, Miranda MF, Fernandes MJ, Rodrigues AM, de Almeida AC, Covolan L. Effects of A1 receptor agonist/antagonist on spontaneous seizures in pilocarpine-induced epileptic rats. Epilepsy & Behavior. 2016 Aug 1;61:168–73.

44. Anzellotti F, Capasso M, Frazzini V, Onofrj M. Olanzapine‐ related repetitive focal seizures with lingual dystonia. Epileptic Disorders. 2016 Mar;18(1):83–6.

45. Arndt DH, Goodkin HP, Giza CC. Early posttraumatic seizures in the pediatric population. Journal of child neurology. 2016 Jan;31(1):46–56.

46. Jabbehdari S, Rahimian E, Jafari N, Sanii S, Khayatzadeh Kakhki S, Nejad Biglari H. The Clinical Features and Diagnosis of Metachromatic Leukodystrophy: A Case Series of Iranian Pediatric Patients. Iran J Child Neurol. Summer 2015;9(3):57–61.

47. Warren PP, Bebin EM, Nabors LB, Szaflarski JP. The use of cannabidiol for seizure management in patients with brain tumor-related epilepsy. Neurocase. 2017 Nov 2;23(5-6):287–91.

48. Artan NS. EEG analysis via multiscale Lempel-Ziv complexity for seizure detection. In2016 38th Annual International Conference of the IEEE Engineering in Medicine and Biology Society (EMBC) 2016 Aug 16 x(pp. 4535–4538). IEEE.

49. Asadi-Pooya AA, Nei M, Sharan A, Sperling MR. Historical risk factors associated with seizure outcome after surgery for drug-resistant mesial temporal lobe epilepsy. World neurosurgery. 2016 May 1;89:78–83.

50. Bergamo S, Parata F, Nosadini M, Boniver C, Toldo I, Suppiej A, Vecchi M, Amigoni A, Da Dalt L, Zanconato S, Perilongo G. Children with convulsive epileptic seizures presenting to Padua pediatric emergency department: the first retrospective population-based descriptive study in an Italian Health District. Journal of child neurology. 2015 Mar;30(3):289–95.

51. Dabscheck G, Prabhu SP, Manley PE, Goumnerova L, Ullrich NJ. Risk of seizures in children with tectal gliomas. Epilepsia. 2015 Sep;56(9):e139–42.

52. Merritt. Neurology. 10th ed. LP Rowland Lippincott Williams & Wilkins eds. Philadelphia. 2000.(8)

53. Glass HC, Numis AL, Gano D, Bali V, Rogers EE. Outcomes after acute symptomatic seizures in children admitted to a neonatal neurocritical care service. Pediatric neurology. 2018 Jul 1;84:39–45.

54. Valente KD, Alessi R, Vincentiis S, Dos Santos B, Rzezak P. Risk factors for diagnostic delay in psychogenic nonepileptic seizures among children and adolescents. Pediatric neurology. 2017 Feb 1;67:71–7.

